# Metaplastic-EEG: Continuous Training on Brain-Signals

**DOI:** 10.1101/2024.05.29.24308178

**Authors:** Isabelle Aguilar, Thomas Bersani--Veroni, Luis Fernando Herbozo Contreras, Armin Nikpour, Damien Querlioz, Omid Kavehei

## Abstract

Deep learning approaches promise viable solutions for detecting epileptic seizures in a reliable, generalisable and potentially real-time. To apply such techniques in clinical settings, where they can be used with long-term recordings or applied to a continuous stream of incoming datasets, these algorithms should adopt a continual learning ability that allows the agent to acquire and adapt from additional knowledge streamed over its lifespan. Unfortunately, traditional sequential learning can initiate catastrophic forgetting, in which the model loses previously learned information while accumulating new knowledge. Metaplasticity has emerged as a potential technique to provide longer-term stability pertaining to the learning performance for multiple datastream sets, thus enabling a meta-learning capability in artificial learning machines and algorithms. In this paper, we use these biologic-inspired metaplasticity techniques to develop stable learning cycles when we expose it to multiple sets of EEG (electroencephalogram) data for seizure detection. In this feasibility study, adding metaplastic synapses enhances detection accuracy relative to traditional baseline learning. Considering the meta-learning approach demonstrated in this paper, metaplastic binarized neural networks (BNNs) demonstrate improvement (6-7%) in seizure detection performance metrics, with reported accuracies and ROC-AUC values over 70%. Metaplasticity in practice with machine learning holds the potential to provide an adaptable, patient-specific epileptic seizure tracking method for real-world dynamics.

## 1. Introduction

Epilepsy affects 50 million people globally, and each year kills an estimated 125,000 persons [1, 2]. On top of an increased death rate, individuals afflicted with epilepsy often face challenges in leading a normal life, encountering a spectrum of issues. These range from medical conditions such as memory loss or a higher propensity for injuries to legal restrictions like a driving ban and extend to social stigmatization like being ostracized due to unfounded fears of contagion [3]. While a significant proportion of epileptic people can be cured either by surgery or medication, some will have to live with occurring seizures their whole lives. Technical solutions to mitigate these seizures, like deep brain stimulation, are being developed but far from readily available [4].

Real-time tracking and forecasting of these seizures are essential for these solutions. Additionally, they would benefit patients by allowing individuals to brace themselves accordingly before the crisis happens. Neurology has advanced to a point where these seizures can be reliably tracked with bio-potential measures like electroencephalogram (EEG) and machine learning algorithms [5, 6].

The existing methods for real-time seizure detection and forecasting require a lot of computational resources and are trained on stationary data, making them unsuitable for implantable, on-the-go adaptive applications. In an implantable context, the algorithms face two major challenges. Firstly, the biomarkers and EEG signatures exhibit high variability between patients and can change over time [7], necessitating a lifelong learning approach for the algorithms. Secondly, these algorithms need to be sparse in power consumption as implantable devices are highly constrained in energy storage. Recent work focused on weak self-supervision to tackle the adaptation issue [8] but did not involve continuous low-power methods. In this paper, we apply a continual learning method for seizure detection.

In lifelong learning approaches, a common dilemma that is faced is catastrophic forgetting, in which new learning interferes with previous learning in sequential training sets [9]. Many methods to address this phenomenon are inspired by the biological networks of our brains. In particular, concepts that describe the plasticity (or adaptive ability) of neural synapses are of interest because they can encode new information while maintaining stability and retention of past memories and experiences [10].

One specific method of interest is metaplasticity, which can be described as the plasticity of plastic synapses [11]. The principles of metaplasticity that have originated in the neuroscience field are potential applications to artificial neural networks (ANNs). Rather than assuming the synapses of the brain are either weak or strong, metaplasticity proposes that multiple mechanisms can influence the degree of synaptic plasticity. Along those lines, previous metaplastic approaches that influence weights and their consolidation in ANNs (such as elastic weight consolidation) have been found to improve recall of old tasks after sequential learning, although none have been adapted into a low-power capacity [12, 13, 14].

## 2. Background

Using electrophysiological data, from surface EEG to epidural and subdural electrocorticogram (ECoG) to endovascular ECoG, have been playing an important role in framing the current and potential solutions for seizure detection in epilepsy. While in this paper, we focus on the EEG-type data for the sake of simplicity and reproducibility of our analysis, our method can be expanded to any electrophysiological data. Seizure analysis with electroencephalogram-type data encompasses seizure detection, where a model tries to detect and/or predict if a patient is experiencing or going to experience a seizure. In literature, both problems have been extensively studied without much attention to the real-time requirements of the potential solutions, limitations of medical recording devices, particularly implantables, and lack of a solution for personalization of analysis. Solutions have been highly skewed towards cloud-based approaches or central training solutions, while we argue that medical-edge-AI with sufficient capability for training could play a major role in personalization.

The field has progressively transitioned from handcrafted expert features [15, 16] to complex machine learning approaches with SVM, random forests, and ANNs [17, 18, 19, 20, 5]. The main goal of these research papers was to find and exhibit the best-performing model on available datasets. As state-of-the-art performance crossed thresholds conducive to real-world testing, some focused on integrating these models into implantable devices [21, 22, 23].

Most implantable-focused research aims at developing low-power architectures to optimize performance on available datasets. However, they do not tackle the variability in EEG signatures between patients and their adaptability to change due to aging or illness [7]. Moreover, only a few implantable-oriented implementations tackle the issue of memory constraints [8] both for the model and the data seen. Indeed, implantable hardware cannot embark on an extreme memory-intensive model and cannot store endless EEG recordings from which to learn. Thus, an implantable implementation needs a model that can adapt to changes in input data distribution and the ability to learn from a stream of data rather than a fixed dataset.

EEG-type data is not infinitely storable in a real-life implantable setting, and the model cannot be re-trained on old data. The streaming nature of real-life data creates challenges, including catastrophic forgetting [9] and patient-specific EEG signature drift that classic cloud-based machine-learning training cannot tackle.

This need for adaptation can be solved using continual learning approaches. These techniques are subdivided into three categories: some models adapt their architecture throughout the learning process [24, 25], others rehearse past seen examples or sampled examples [26, 27], while other models employ penalty and regularization techniques to retain important features while discarding unimportant ones [28, 12, 29]. Some of these techniques are brain-inspired to reduce the power and memory usage of continual learning heavily [14], including metaplasticity, which is examined and applied here (see Fig. 1. These techniques, sitting between low-power and life-long learning, are the ideal adaptation to EEG analysis to tackle the aforementioned challenges in an implantable setting and what we will investigate in the present report.

**Figure 1:**
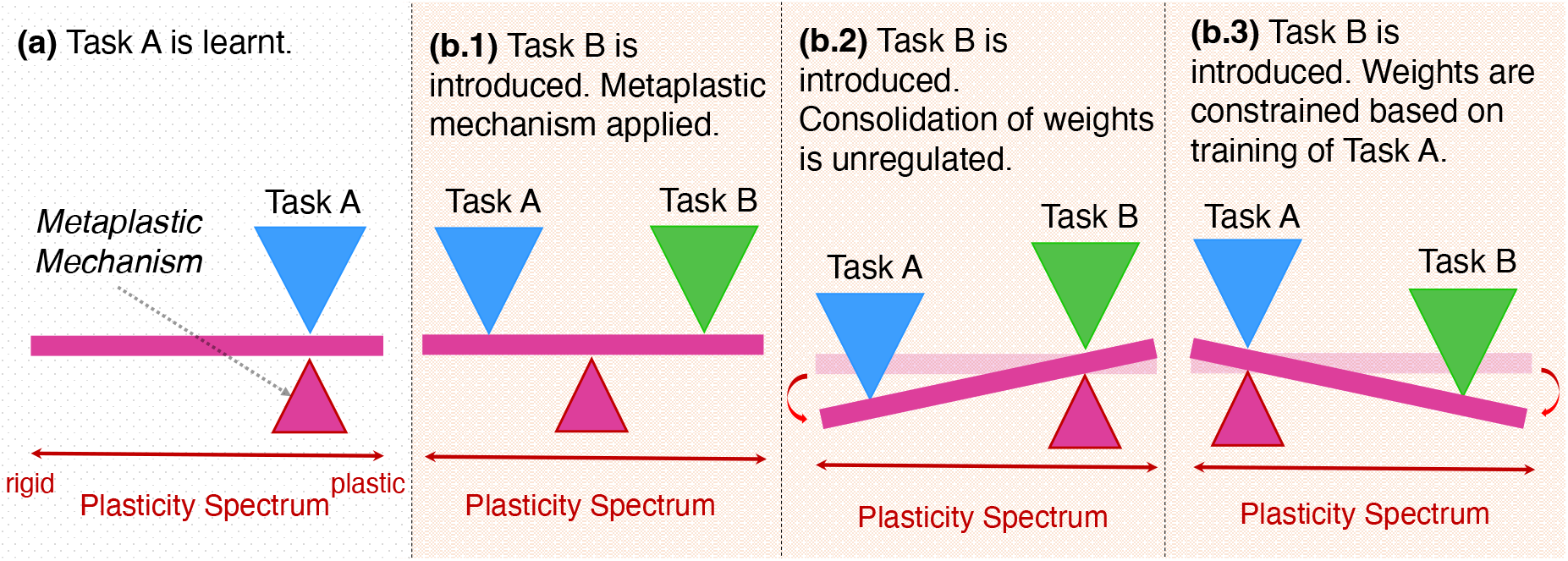
Representation of metaplasticity. Consider a seesaw, where the fulcrum (acting as a metaplastic mechanism) can move along the beam, determining the plasticity of a system. a) Task A is introduced to the system and is represented by a blue triangle. b) Task B is introduced into the system. b.1) Metaplastic mechanism has shifted along the plasticity spectrum to reach a balance between rigid/plastic synaptic states to maintain performance in both tasks and perform with low error. b.2) With unregulated consolidation of weights, Task B is learned at the risk of losing information learned from Task A. Catastrophic forgetting occurs. b.3) On the other hand, if weights are too constricted, the network is unable to retain knowledge for Task B.

Several EEG datasets are freely available for seizure detection and prediction. The most frequently used datasets originate from the Children Hospital of Boston - MIT [30], UPenn and Mayo Clinic [31], and Temple University Hospital (TUH) [32]. Among these datasets, only the TUH dataset has long enough records and a significant number of patients to enable a study on continual learning [33].

While seizure detection and seizure prediction are both achievable on this dataset, it is not long enough to perform seizure prediction in a continual learning fashion. We thus limited our work here to seizure detection.

## 3. Methods

### 3.1. Data Preparation

The Temple University Hospital (TUH) EEG Seizure Corpus v1.5.4, is used for training and testing our model. The TUH dataset consists of recordings of 642 patients at 250Hz. The training set encompasses 592 patients, and the development set is 50. We use the development set as a testing set here, resulting in a training set of 752 hours and a testing set of 47 hours.

Initially, the TUH dataset possessed imbalanced labels between seizure and background. To tackle this issue, we apply a technique previously used in the literature [20]. We generate more ictal segments by sampling with an overlap of the original ictal segments. As a result, we successfully reduce the imbalance to a 30%/70% repartition between ictal and background segments.

### 3.2. Preprocessing

Metaplasticity, which is visualized in the previous section and encompasses our main approach to continual learning, is a technique primarily developed and tested in an image-processing context. We thus need to transform the EEG data, a multi-channel time series, into an image-like input. Fourier transforms and wavelets have been successfully used in the literature on EEG data [20, 34]. In this work, the log-magnitude Short-Time Fourier Transform (STFT, see Equ. 1*a*) is used to transform a two dimensional time-series into a time-frequency tensor. To perform the Fourier transform, we use 12 seconds data points, a von Hann window (see Equ. 1*b*) of 1 second, and an overlap of 50%. We then recover a tensor of dimension (channel *×* time *×* frequency).

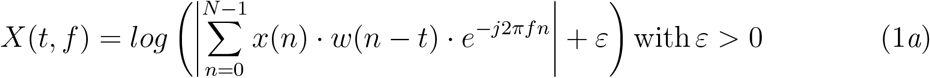

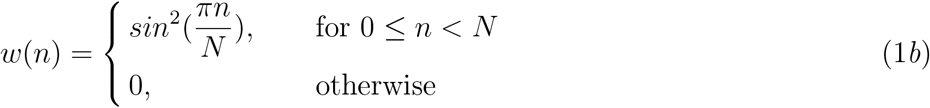

Like most EEG recordings, the TUH dataset is subject to artifacts from the utility frequency of power lines (see Fig. 2 before artifact rejection). Therefore, the frequencies around 60Hz*±*5Hz are removed, as well as the DC component (at 0 Hz) in respect to the minimum resolvable frequency (0.08Hz). We then average the tri-dimensional tensor over the channel dimension to collapse it into a bi-dimensional tensor suited for our metaplasticity application (see Fig. 2).

**Figure 2:**
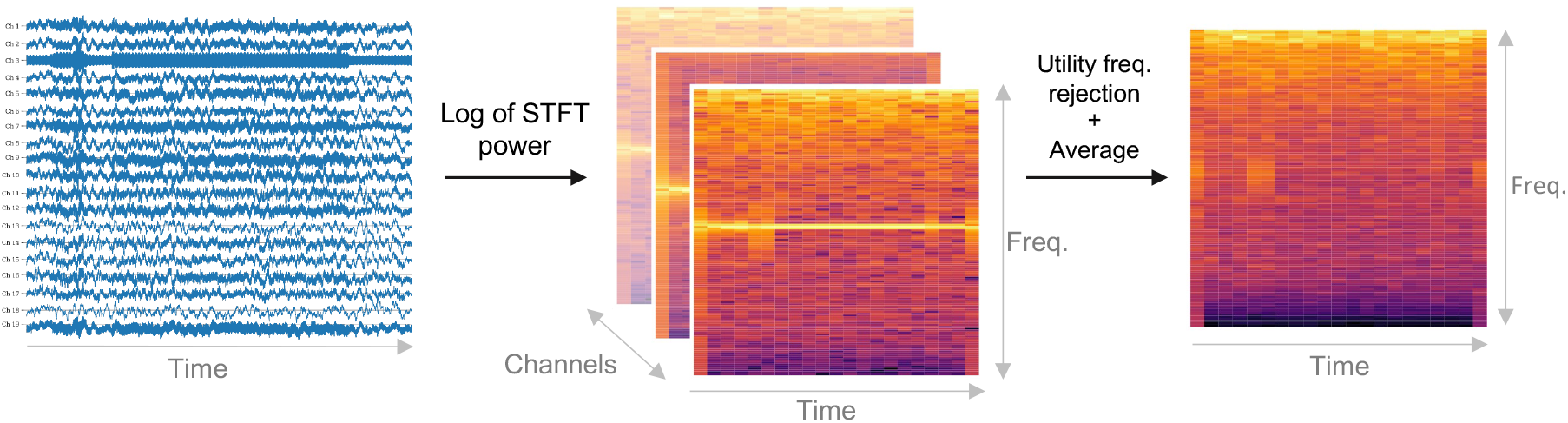
Preprocessing steps. Starting from a time series to an image-like data point through STFT and filtering.

### 3.3. Binarized Neural Network

Binarized neural networks (BNNs) are developed to reduce the memory and energy usage of deep neural networks. These systems are competitive with full-precision networks in terms of performance on a range of tasks [35, 36, 37]. This is particularly attractive in the context of implantable devices, as it addresses memory constraints and the necessity of energy efficiency.

These neural networks are unique in that their activation functions and weights are constrained to +1,*−*1. They learn through back-propagation on a real weight that will accumulate the gradient updates. This real-valued weight is said to be hidden because the inference stage is done with the sign of the hidden weight (termed the binary weight) as is the gradient update computation. The bit-wise operations of BNNs provide a more compact model that is ideal for the deployment of edge devices for real-time, low-power applications. By offloading computations from the cloud to the edge, we not only enhance performance and responsiveness but also improve data privacy and security.

In terms of its architecture, the network used as a baseline in our work a two-hidden-layer-wide BNN with 4096 neurons on each hidden layer. Batch-normalization is used after every layer but the last one, in order to yield a more stable training and better performances [35, 38]. The hidden weights of the network are initialized with a uniform distribution over [*−*0.5, 0.5].

### 3.4. Metaplastic Learning

Synaptic metaplasticity [14] is developed to tackle catastrophic forgetting in a highly-constrained power setting. The absolute value of the hidden weight, or binary weight, is interpreted as the importance of the synapse, denoted by the network, during the learning process. In this approach, the higher the absolute value of the hidden weight, the more difficult it should be to flip its sign, as the network grew this magnitude toward a sign repeatedly during the training.

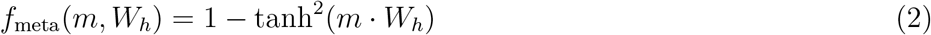

Laborieux et al. [14] introduced a metaplastic function *f*_*meta*_ defined in Equ. 2, where *m* is a hyperparameter and *W*_*h*_ is the hidden weight. This function *f*_*meta*_ will tone down the gradient update on the hidden weight if it diminishes its absolute value, as defined in algorithm 1. The parameter *m* controls how well the network remembers past seen examples, and how quick the network weights will be consolidated, i.e. desensitized to new data inputs. Fig. 3 describes this approach.

**Figure 3:**
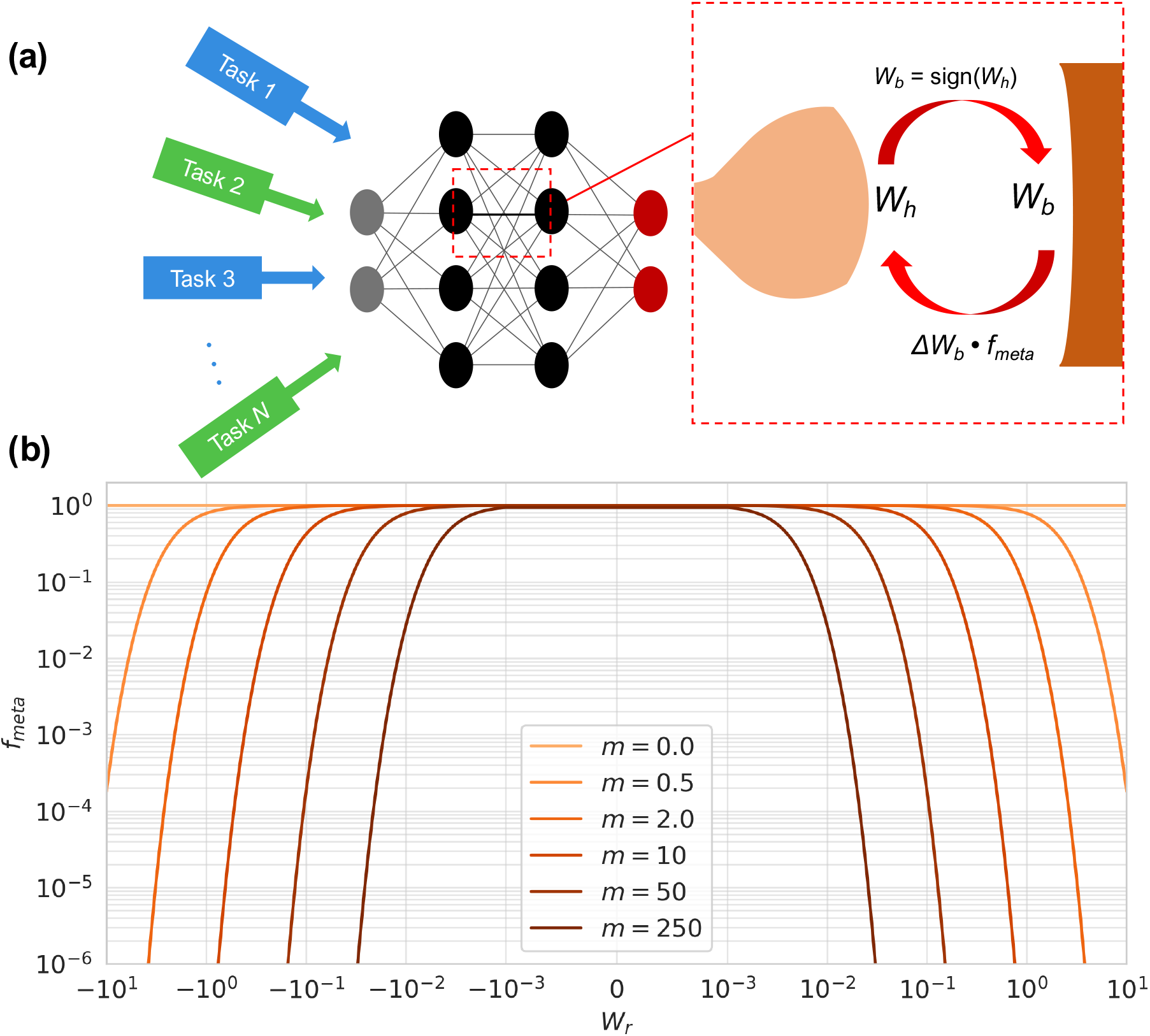
Metaplastic binarized neural network (BNN). a) In a BNN, metaplasticity is applied by considering the hidden real weights as a metaplastic component which influences a metaplastic function, *f*_*meta*_, which affects gradient updates in the network. b) The function is chosen so that an increase in the magnitude of the hidden value makes it more difficult to switch synaptic states. As the metaplastic hyperparameter, *m*, increases, the synapses of the network are more rigid.

In regards to additional parameters, the learning rate used in this work is set to *η* = 10^*−*4^. The Adam betas (*β*_1_, *β*_2_) are set to 0.9 and 0.999 respectively, the default values of PyTorch v.1.12.1.

### 3.5. Continual Data Stream

To assess how well the proposed model could behave on a real-life continual learning environment like AURA (an efficient semi-supervised model that trains a seizure forecasting task with unlabeled, real-time data and autonomously generated labels) [8], we first propose to test its performances on a stream learning setting to place the model in a close to implantable data-feed. The streaming task consists of training and testing the model on one task (in our case, epileptic seizure detection), in which the network learns sequentially on subsets of the data that are revealed one at a time. This stream environment reflects a realistic situation of instantaneous temporal data flow in an implantable EEG device, where previous information is not stored, and we do not have access to subsequent data signals. The TUH training dataset is randomly divided into 300 subsets that are sequentially fed to the model for 20 epochs each. We test the model every 5 subsets (for a total of 60 times) on the test dataset to see if it can generalize by remembering data seen only once during the training. Due to the limited training data available, in order to prevent overfitting, the BNN model was simplified to 2048 neurons on each hidden layer. In addition, other mitigation methods were applied: L2 regularization and data augmentation via Gaussian noise addition at different time points of the TUH dataset [39].

#### Algorithm 1

Metaplastic update on a single hidden weight. **W**_**h**_ is the vector of hidden weights, and *W*_*h*_ denotes one component (the same rule is applied for other vectors), **U**_**W**_ is the weight update yielded by a classic Adam gradient descent algorithm, (*x,y*) is a batch of labelled training data, m is the metaplasticity parameter, and *η* is the learning rate.

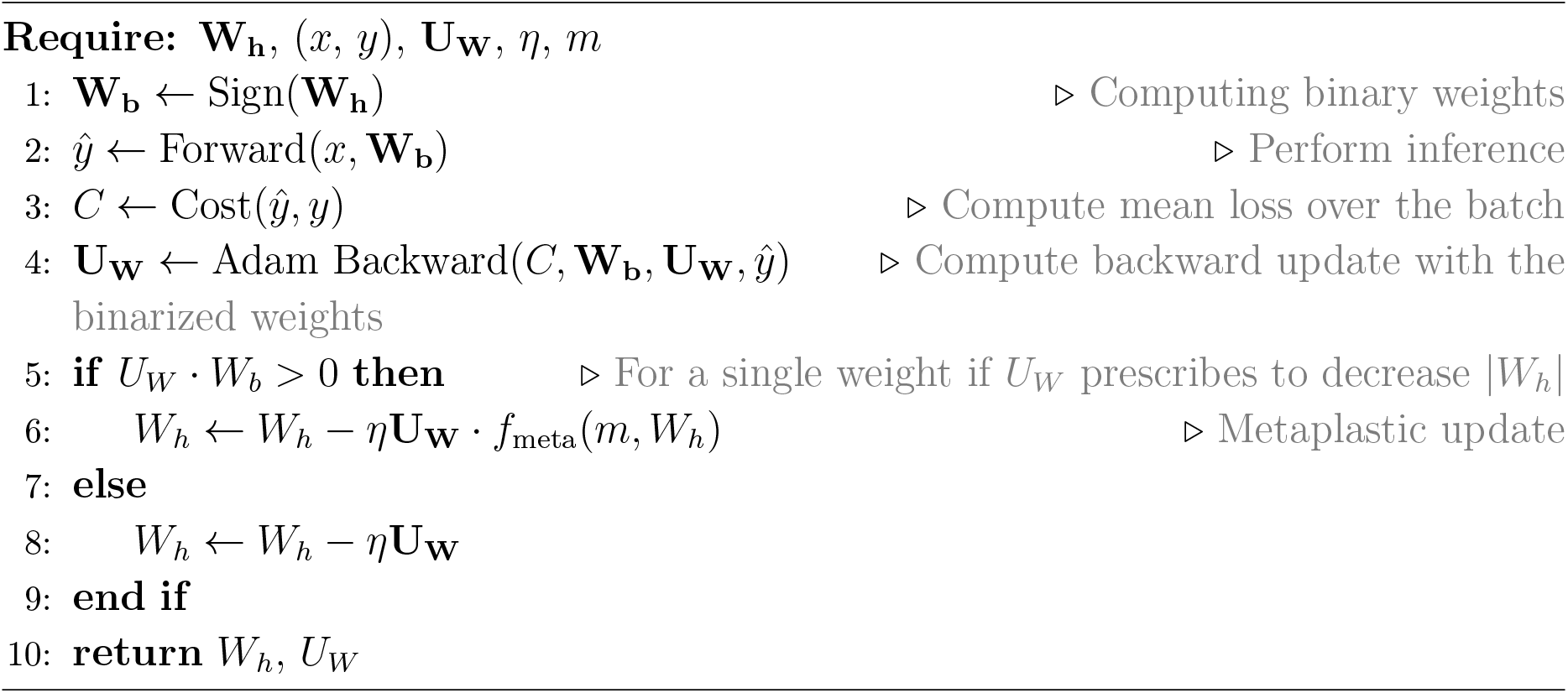

### 3.6. Adaptation to Seizure Pattern Changes

We then assess the performances of the proposed method on different EEG frequency signatures to see if the network can adapt to significant changes in seizure patterns. Amongst different patients, seizure patterns often differ. Even within a single patient, these patterns can change over the course of a lifetime due to aging or illness.

For this purpose, five synthetic EEG datasets are generated with different seizure signatures. For a given synthetic dataset 𝒟, we pick a random permutation of pixels ℱ_*𝒟*_ such that every element *x*_*𝒟,i*_ of 𝒟 is equal to *x*_*𝒟,i*_ = ℱ_*𝒟*_ (*x*_*TUH,i*_), the permutation of the element of same index in the original dataset. Amid the five datasets used, one is the original TUH while the other four are synthetic.

The model trains sequentially on the five datasets for 20 epochs each, or 100 in total. We evaluate its performance during the whole training on the test set of each synthetic dataset.

### 3.7. Performance Evaluation

#### 3.7.1. Seizure Detection Performance Metrics

To evaluate the relevance of this proof of concept, we are set in a seizure detection problem. The classic performance metrics of this task are the AUC-ROC Score (equ. 4) [19] and accuracy (equ. 3). Accuracy is defined as:

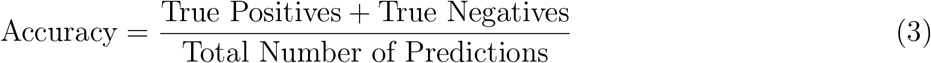

This metric quantifies the proportion of correct predictions made by the model over the total number of instances evaluated. The AUC-ROC score is defined as:

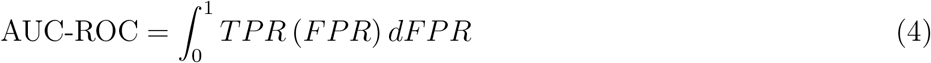

where *TPR* is the True Positive Rate, and *FPR* is the False Positive Rate. ROCs (receiver operating curves) quantify a model’s ability to discriminate between positive and negative classes. The AUC score ranges from 0 to 1, with 1 indicating perfect discrimination and 0.5 indicating an algorithm has no better performance than random chance.

Tracked together, these two metrics enable a global and balanced evaluation of the model performances in seizure detection. In this data-imbalanced task, the cost of a false-negative is higher than that of a false-positive.

#### 3.7.2. Continual Learning Tasks

To track how well the model performs in a continual setting, we visualize the classic seizure detection metrics defined in section 3.7.1 throughout the whole training; we also use time averages and final points of these metrics.

For a multi-task setting as in 3.6 we define and use the time-average 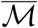 of a metric ℳ as the average of the metric over the learned tasks at the epoch step as seen in Equ. 5, where *T*_*max*_(*epoch*) is the latest learned task at a given epoch.

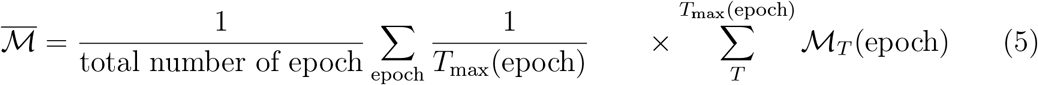

For a stream learning setting as defined in section 3.5, we use the final metric value to assess if the model has a global understanding of the dataset learned.

## 4. Results

### 4.1. Streamed Data and Continual Learning

All results are averaged over six training runs. The dataset is reshuffled between every training.

Fig. 4 presents the model’s performance on the whole test set while learning on a sequential subset of TUH for different values of the *m* parameter. The results of training a BNN on the full dataset for 20 epochs with the same *m* values are also reported as a reference. The vanilla BNN’s performances degrade after the first 50 subsets learned to values below 0.75 for both AUC and Accuracy. Overall, the performance of the vanilla BNN is also noticeably unstable. In comparison, the metaplastic network (*m* = 30) shows AUC and Accuracy values ranging between [0.73-0.78], which is of greater value than the performance metrics of the network trained on the streamed dataset with zero metaplasticity.

**Figure 4:**
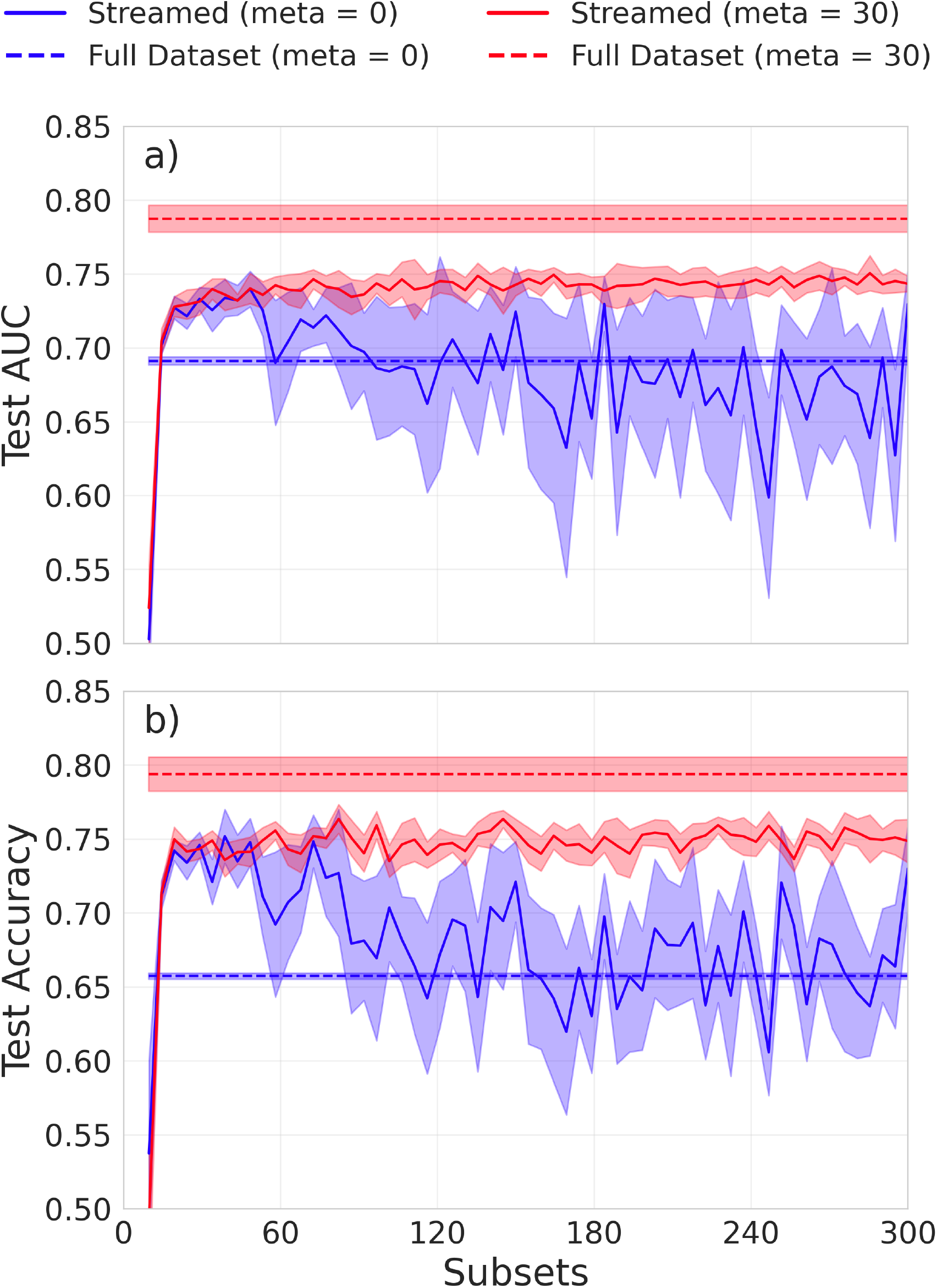
Test dataset metrics on continual learning of seizure detection on the TUH EEG dataset. a) measures AUC and b) measures Accuracy. Averaged over six runs, the solid curves are the results of training on a vanilla and a metaplastic BNN model sequentially on a tiny batch of data for 20 epochs each. The shaded area represents one standard deviation. The dashed lines represent the result of the same type of model trained on the whole dataset for 20 epochs (i.e., no streaming AKA conventional learning).

In Table 1, we present the last values of the test metrics for this experiment. We observe an overall improvement of 6% and 10% of the ROC-AUC and Accuracy, respectively, when the metaplastic BNN is used compared to a vanilla network.

**Table 1:**
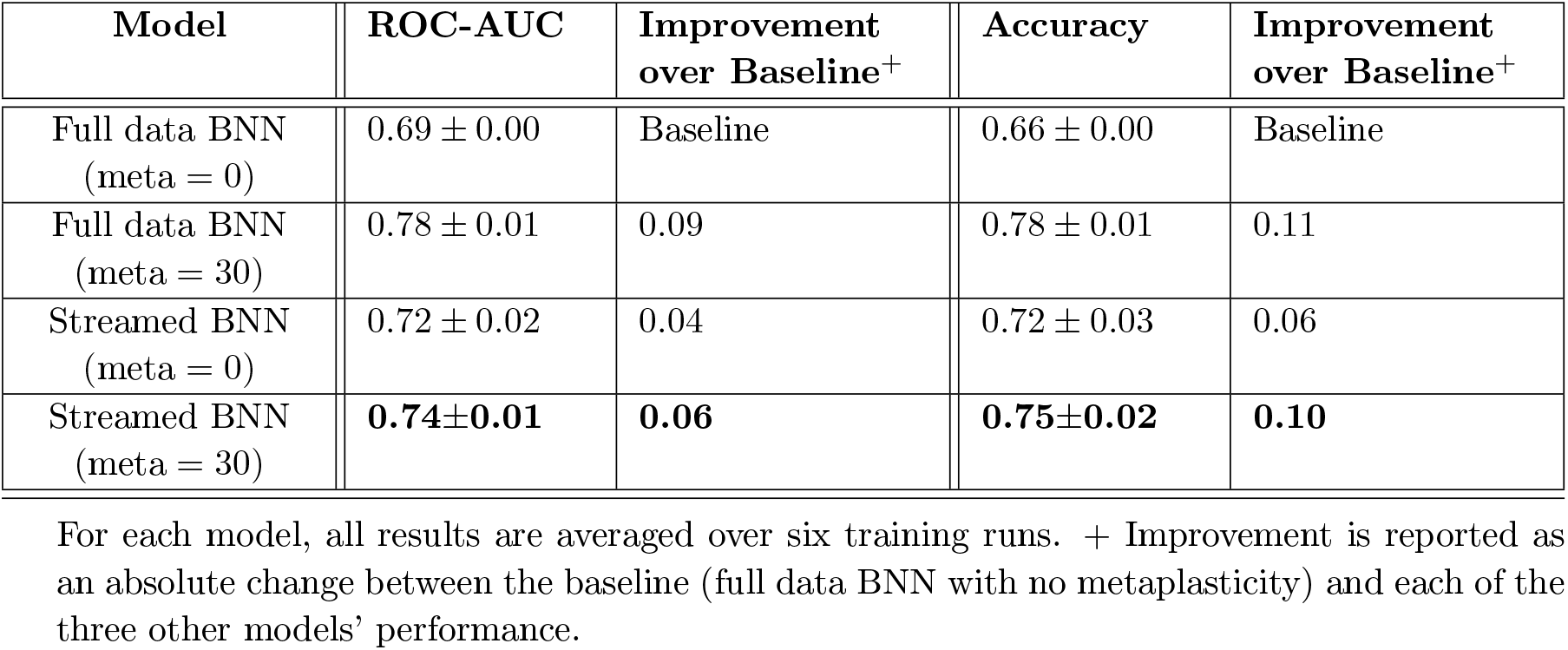
Stream-learning performance test metrics versus conventional full-data feed in BNNs with and without metaplasticity. Test ROC-AUC and Accuracy values are presented along with their standard deviations. For streamed BNN models, final test metrics are reported (see Sec. 3.7.2 for further details).

### 4.2. Synthetic Data

All results are calculated on the test dataset on five different training runs. Between each training session, every synthetic dataset is re-generated by picking new random permutations, and the order of presentation of datasets is randomized.

Fig. 5 presents the test accuracy and ROC-AUC of each dataset during their sequential learning for a classic BNN (*m* = 0) and metaplastic BNNs with an *m* parameter of 30 and 60, with 4096 neurons on each layer. The vanilla BNN forgets a task learned as quickly as it learns the new one; see Fig. 5(a), depicting a clear case of catastrophic forgetting. When metaplasticity is introduced, the model maintains its performance in a more proficient manner in comparison to the vanilla network.

**Figure 5:**
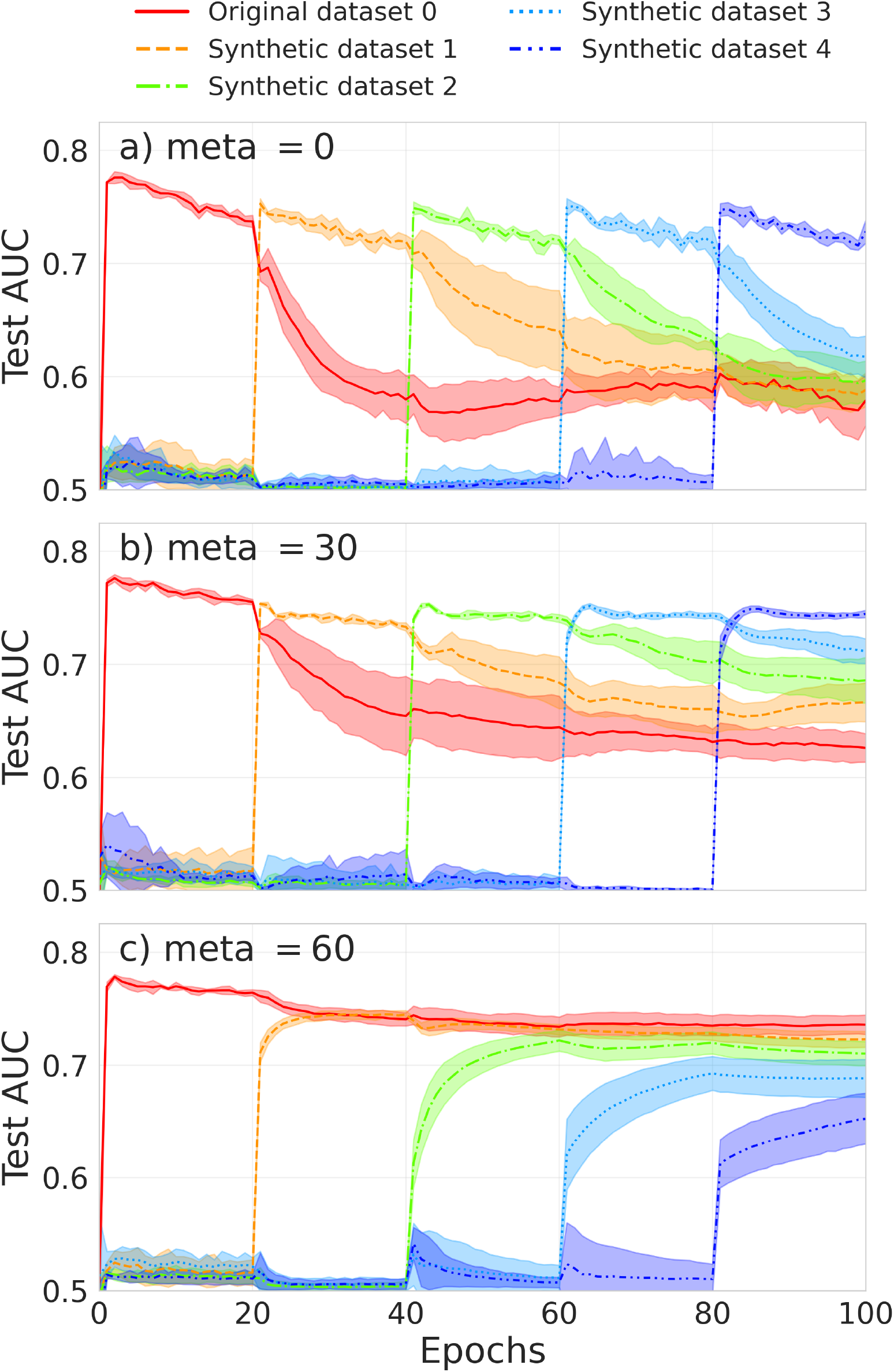
a-c) Binarized neural network learning five datasets of differing seizure signatures sequentially for several values of the metaplastic parameter *m*. a) *m* = 0 corresponds to a vanilla BNN, b) *m* = 30, c) *m* = 60. Curves are averaged over five runs and shadows correspond to one standard deviation.

The two metaplastic models show a recall of the previously seen tasks. In Fig. 5(b), the model exhibits a slower forgetting of the task learned, and reaches both peak accuracy and ROC-AUC that are the same as the vanilla ones. Fig. 5(c) displays the models maintaining their performances throughout the whole training but failing to reach the earlier peak performance seen by the two other networks. It exhibits the trade-off between being able to recall old material and being able to learn new material. This trade-off is ultimately imposed by the size of the network as more synaptic connections enable more storage for information. Table 2 presents the time-averaged metrics as defined in 3.7.2 for different *m* parameters and a size of 4096 neurons for each hidden layer.

**Table 2:** Performance metrics in seizure detection results obtained with the TUH EEG dataset. The training was executed five times, and time-average results for test ROC-AUC and Accuracy with standard deviations are reported.

| Model | ROC-AUC | Improvement over baseline<sup>+</sup> | Accuracy | Improvement over baseline<sup>+</sup> |
| --- | --- | --- | --- | --- |
| Vanilla BNN (meta = 0) | $0.67 \pm 0.01$ | Baseline | $0.68 \pm 0.00$ | Baseline |
| BNN (meta = 30) | $0.71 \pm 0.01$ | 0.04 | $0.71 \pm 0.00$ | 0.03 |
| BNN (meta = 60) | **$0.73 \pm 0.01$** | **0.07** | **$0.73 \pm 0.00$** | **0.05** |

All models contain 4,096 neurons per hidden layer. + Improvement is reported as an absolute change between the baseline (vanilla BNN with no metaplasticity) and each of the two other models' performance.

## 5. Discussion

Synaptic metaplasticity in BNNs present an opportunity to realize a low-power, easy-to-implement, and relatively portable alternative to conventional networks. We tested the model on a stream of EEG time-series data to evaluate the its performance for future bionic use with a need for real-time response. We then tested the system on different signal features, specifically on its adaptability to high amplitude changes in the frequency domain. Both tests resulted in a validation of the network performance. However, we also observed a diminishing return phenomenon in which the increase of the metaplastic parameter beyond a certain value (around 60) reduces the network’s ability to recall learned information. Establishing the optimal value for metaplasticity to maintain this balance is ultimately imposed by the network size, as more synaptic connections will enable more information storage. Overall, the model consistently performed better when metaplasticity was introduced, with an average increase of 7% in absolute value with a metaplasticity value of 30. This result gives us confidence in using this technique for future work.

While there is an appreciation for real-time detection/prediction of medical abnormalities in time-series data [40], its prevalence in the continual learning literature is not well understood or discussed. The significance of such continual learning and adaptive systems is particularly significant for brain-machine interfacing (BMI), where the current impractical combination of raw-data telemetry bandwidth and power requirements creates massive hurdles for implantable BMIs. Due to this restraint, we faced limitations in comparing our results to state-of-the-art benchmarks.

As the existing benchmarks are done on stationary data, with no energy and time dependence issues, a comparison at face value would not be meaningful. However, it is significant to note that our simple fully connected binary classifier model reaches the same neighborhood of 0.80 ROC-AUC and Accuracy that was reported in the literature for a network trained on stationary data with full precision and fully connected networks [41]. This also gives us confidence on the viability of the used technique, the fully connected and state-of-the-art performances relied on better feature extractors that were not necessary for our system. Ultimately, the metaplastic BNN imposes a simplistic, biologically-inspired model that is able to compete with the performance of fully connected systems that require more computational resources. This method has shown usefulness in the case of ictal EEG, in which signals are heterogeneous across a population but stereotyped in individuals.

Further studies may benefit from the application of automatic feature extractors like a convolutional neural networks (CNN) or recurrent neural networks (RNN). Additionally, the use of longer data series that can conduct a deeper analysis on our system is ideal, although a lack of such public records are a common barrier in biomedical research.

## Data Availability

This paper employs a publicly accessible Temple University Hospital EEG Data Corpus. This corpus can be found via the Neural Engineering Data Consortium.

## Acknowledgements

The authors acknowledge the support from the Australian Research Council under Project DP230100019. Isabelle Aguilar would like to express gratitude for the generous assistance and support the Australian Government’s Research Training Program (RTP) has provided. Luis Fernando Herbozo Contreras would like to acknowledge the partial support of the Faculty of Engineering Research Scholarship provided by the University of Sydney. Damien Querlioz acknowledges support from the EIC Pathfinder METASPIN grant (reference: 101098651).

## Code Availability

Codes will be made accessible upon request to the corresponding author.

## Conflicts of Interest

All authors declare that they have no conflicts of interest to disclose.

## Notes

### Competing Interest Statement

The authors have declared no competing interest.

### Author Declarations

This study has used the publicly available Temple University Hospital EEG Data Corpus dataset, provided at this link: https://isip.piconepress.com/projects/tuh_eeg/html/downloads.shtml

